# Increased sensitivity of detecting variation in malaria endemicity by parasite density measurement in large surveys

**DOI:** 10.64898/2026.10.07.26364850

**Authors:** Wellington Oyibo, Alexandra M. Blair, Oladosu Oladipo, Nnenna Ogbulafor, Rita Omohode Urude, Ginika Onwuachusi, Chinonye Anabike, Godwin Ntadom, Chukwu Okoronkwo, Perpetua Uhomoibhi, Nelson Eze, Olusola Oresanya, Aminu Mahmoud, Onyebuchi Okoro, Bolaji Aduagba, Simon Ijezie, Festus Okoh, Issah Kawu, Lynda Ozor, Ibrahim Maikore, Kolawole Maxwell, Sonachi Ezeiru, Ernest Nwokolo, Chioma Amajoh, Mohammed Audu, Olugbenga Mokuolu, Zugbema Biyama, Temitope Adeyoju, Nwohu Obike, Joseph Ilelaboye, Ayebatari Lawson, David J. Conway

**Author notes:** Corresponding authors: Prof Wellington Oyibo; Prof David Conway.

## Abstract

**Introduction:** The continuing high and inequitable global malaria burden requires improved methods to understand trends and target control. Current methods often involve infection prevalence surveys, but additional parasitological measures could increase precision in estimating infection burdens of communities.

**Methods:** Focusing on the country with most malaria globally, examinations of slides from 10,651 children under five sampled in the 2021 national Malaria Indicator Survey in Nigeria were performed to measure asexual parasite densities in infected children and discriminate sexual stages and species. Variation in these indices among the six different geopolitical zones and 37 individual states including the Federal Capital Territory were compared with the variation indicated by infection prevalence alone.

**Results:** Among the six major geopolitical zones, only one had significantly higher asexual parasite prevalence than others, but parasite densities within infected children revealed more significant variation among the different zones. Comparing results for 2021 with previous data from 2018, one zone showed lower asexual parasite prevalence and two had lower sexual parasite prevalence, but three had higher asexual parasite densities in infected children. Variation in parasite densities among individual states also highlighted significant geographical and temporal differences, extending the detection of heterogeneity within each geopolitical zone. Overall trends were driven by the predominant parasite species *Plasmodium falciparum* as prevalence and density of the other species were much lower.

**Conclusions:** Findings indicate that understanding complex patterns of geographical and temporal variation is enhanced by including asexual parasite density measurement, providing opportunities for more precise targeting of malaria control within high burden countries.

**Key Messages of Article:** *What is already known on this topic:*

- Heterogeneity in malaria infection burden among endemic communities is commonly assessed by surveying parasite prevalence.
- Previous analysis of blood stage malaria parasite densities suggested that incorporation of this variable might contribute to a more thorough assessment.

*What this study adds:*

- Analysis of the 2021 national survey in Nigeria shows that incorporation of parasite density data enhances detection of heterogeneity, among major geopolitical zones and individual states.
- Comparison with 2018 data identifies variation over time more sensitively, with some communities showing an increase and others a decrease in infection burden.

*How this study might affect research, practice or policy:*

- Given only marginal extra resources needed to obtain the additional microscopy measurements, on top of scoring parasite positivity, these may be recommended to enhance monitoring and targeting of malaria control.

## Introduction

The highly inequitable distribution in global malaria burden is becoming more apparent. A trend of progressive reduction in malaria since 2000 has stalled since 2015, and recent estimates indicate that numbers of cases are increasing, particularly in high-burden countries where populations are growing ^1^. In a contrasting trend, many countries that had low infection burdens have achieved malaria elimination since 2015 ^2^. More than half of all malaria cases and deaths now occur within West Africa, and more than half of these are in Nigeria which bears approximately 30% of the current global malaria burden ^1^.

The level of health inequity within many endemic countries is key to the challenge for malaria epidemiology and control ^1^ ^3^. The most reliable relevant information within many countries has been provided by national Demographic and Health Surveys (DHS) and Malaria Indicator Surveys (MIS) co-ordinated by the DHS Program ^4^. Such surveys that apply rigorous methods enabling effective comparisons among regions within countries are of more importance than ever, but it is increasingly difficult to obtain funding to sustain them ^5^. Securing future support depends on evidence-based analysis of the value of survey measurements, which encourages country-led innovation ^6^.

For malaria surveys, a standard measurement has been the point prevalence of infection in community surveys of children ^7^, but there are limitations to its informativeness. Firstly, it only considers presence or absence of detected infection in each sampled individual, whereas infections have a very wide range of blood parasite densities ^8^. Secondly, parasite prevalence estimates are subject to varying sensitivity depending on the detection method applied ^9^ ^10^, including the standardized method of microscopical detection of parasites on Giemsa-stained blood smears ^11^.

Although most of the population of Nigeria is at risk of malaria infection, the endemic burden varies substantially throughout the country, and understanding this is vital for sub-national tailoring of interventions as now recommended by WHO ^12^. National surveys of children under five in Nigeria using quality-controlled malaria parasite microscopy have been conducted, with prevalence results published for surveys in 2010, 2015, 2018 and 2021^13–16^. These show there was a decrease in overall prevalence of malaria infection in children under five years of age between 2010 and 2015 ^13^ ^14^, but no significant decrease at a national level in the subsequent 2018 or 2021 data ^15^ ^16^. However, detailed analyses reveal major variation in prevalence between different geopolitical zones of the country, as well as variation in the temporal trends ^17^. Some zones and individual states within them that had declines in prevalence between 2010 and 2015 also had further declines in prevalence subsequently, while others had no further decline or showed increases ^17^.

Tracking of malaria trends may benefit from additional measurements, so an initial analysis was performed on densities of parasite infections and presence of parasite sexual transmission stages within infected children, by detailed malaria microscopy of the slides from the 2018 national survey in Nigeria ^18^. This showed that the mean density of parasites in infected children varied throughout the country, and broadly correlated positively with the prevalence of infection, being significantly higher in the northern zones of the country than in the south ^18^. Furthermore, analysis of parasite density data from individual states revealed significant within-zone variation independent from the prevalence measurements, suggesting that the use of both variables together yielded a more complete indication of infection burden.

To investigate the value of extending beyond infection prevalence estimates, the present study investigates parasite density and additional microscopy measurements from slides sampled from children under five years of age across all states of Nigeria in the 2021 Malaria Indicator Survey. Results show that parasite density measurements as well as discrimination of sexual stages increased the resolution of geographical and temporal variation in infection burden. This highlights the potential for programmatic use of these indices to be included for monitoring and targeting malaria control in high burden countries.

## Methods

### Study population and sampling

To assess the prevalence of malaria asexual parasites in children under five years of age in Nigeria in 2021, a nationally representative sample of children aged 6 to 59 months was selected across all 36 states as well as the central Federal Capital Territory (FCT), throughout the six geopolitical zones of the country (Figure 1A). As part of the Nigeria Malaria Indicator Survey (NMIS), trained biomarker laboratory scientists collected blood samples following informed consent from each child’s parent or guardian ^16^. Samples were obtained via finger prick or heel prick (for children aged 6–11 months). Thick and thin blood smears were prepared in the field according to WHO-recommend procedures (Standard Operating Procedure SOP05a) ^19^. Each slide was labeled with a unique barcode and transported to one of 18 regional laboratories within Nigeria for Giemsa staining. Subsequently, stained slides were sent to the African Network for Drug and Diagnostic Innovation (ANDI) Centre of Excellence at the College of Medicine, University of Lagos, for logging, storage, and microscopic examination.

**Figure 1.**
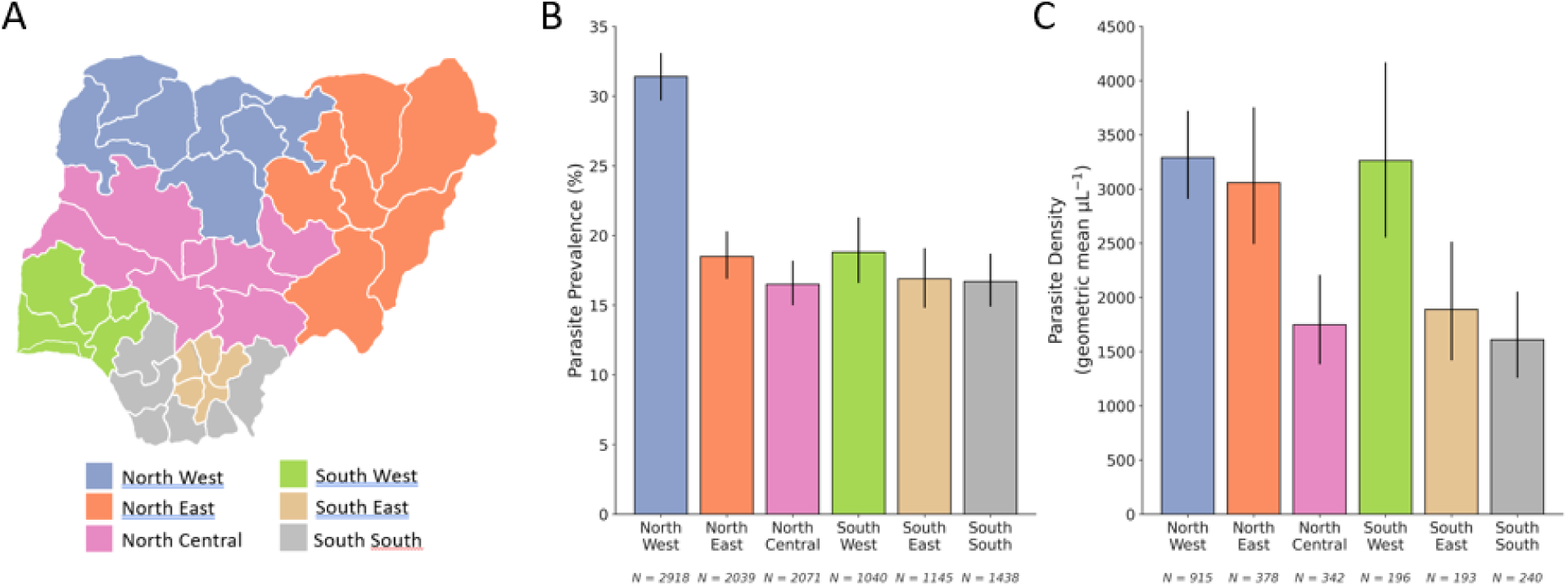
Malaria infection prevalence and asexual parasite density across geopolitical zones in Nigeria in the 2021 national survey. The colour shading on the map of Nigeria shows the individual states that are grouped into six major political zones, and the numbers of slides from the 2021 Malaria Indicator Survey for which full microscopy results were obtained for the analysis here. There was significant variation in prevalence of positive slides across the geopolitical zones (P < 10^-15^) as the North-West zone had a much higher prevalence (31.4 %) than the other five zones. There were differences among the geopolitical zones in the asexual parasite density in positive slides (P = 0.0026), with high geometric mean densities in the North-West (3291 μL^−1^), North-East (3059 μL^−1^) and the South-West (3262 μL^−1^), whereas the remaining three zones each had lower means of <2000 μL^−1^. The indices and numbers are tabulated in Supplementary Table 2.

### Quantification of malaria parasitaemia by microscopy

All slides were independently read by two trained microscopists at the ANDI laboratory, each microscopist recording the presence of asexual malaria parasites, gametocytes, parasite species identification and parasite densities. The results for slides with discordant readings were resolved by a third expert reader. Microscopy adhered to WHO-recommended quality assurance procedures ^20^, with parasite quantification performed by counting numbers against 200 white blood cells (WBCs) or per 500 WBCs when necessary ^21^. Measurement of parasite density for each slide was based on the mean of the two independent readings, and if these differed by more than two-fold a third reading was used to determine the parasite densities by recomputing the mean parasite count with the nearest of any of the initial two readings. Any missing values were handled using complete-case analysis, with records excluded pairwise depending on the variables involved.

### Data Processing and Cleaning

Microscopy data including parasite detection, parasite densities, asexual and sexual stages, and *Plasmodium* species identification were maintained in Excel format and checked for completeness. These data were merged with the individual demographic records from the 2021 NMIS survey ^16^, using the unique slide barcode as the linkage key. Cross-tabulations were conducted to ensure consistency between matched datasets, in which records with unmatched or duplicate barcodes were excluded.

For comparison, data were also analysed that had been derived from examinations of Giemsa-stained malaria blood slides from the 2018 Nigeria Demographic and Health Survey (NDHS) ^15^ that used the same sampling and processing protocols with co-ordination by the DHS and National Malaria Elimination Program. The asexual parasite density and sexual parasite prevalence data in these 2018 survey samples have been separately reported ^18^, and are used here for the comparative analysis.

### Statistical Analysis

Summary statistics were computed to describe the prevalence and density of asexual malaria parasites and presence of gametocyte-stage parasites in all geopolitical zones and individual states. Categorical variables were summarized as frequencies and percentages. Parasite densities, which follow a right-skewed distribution, were summarized using geometric means and 95% confidence intervals, calculated via log transformation and back-transformation. Parasite density values of zero were excluded from log-transformed analyses and geometric mean calculations.

Due to the non-normality of parasite density distributions in blood, non-parametric approaches were used for significance testing. Parasite density values of zero were excluded from log-transformed analyses used to calculate geometric means and corresponding 95% confidence intervals. Non-parametric statistical tests were used to assess differences between different sub-groups, the Mann–Whitney U test for two-group comparisons and the Kruskal–Wallis test for three or more groups. Prevalence estimates were calculated using Wilson confidence intervals, and significance of correlations was examined using Spearman’s rank test. All analyses were performed using R version 4.1.0 or later.

### Patient and public involvement

Patients or the public were not involved in the design, or conduct, or reporting, or dissemination plans of our research.

### Ethical Approval

All samples were collected as part of the 2021 Nigeria Malaria Indicator Survey (NMIS), conducted under the Demographic and Health Surveys (DHS) Programme, the National Malaria Programme and other partners including the National Population Commission of Nigeria. The National Health Research Ethics Committee of Nigeria (NHREC) and the ICF Institutional Review Board approved the survey protocol, as described in the published 2021 NMIS Report ^16^. The London School of Hygiene and Tropical Medicine (LSHTM) Ethics Committee approved the research involving combination of publicly available demographic datasets and the corresponding previously unpublished data on malaria parasite density from the microscopical slide analysis (Project 30513, Review RR/35148).

## Results

### Measurements of malaria parasite density in children under five years of age throughout Nigeria

Full data were obtained for 10651 children under five years of age, after merging the detailed microscopy results including parasite density with demographic variables on the same individuals in the 2021 Nigeria Malaria Indicator Survey. Overall, 21.4 % (2275) of the children were slide positive for malaria parasites. Of these 91.0 % (2071) had only *Plasmodium falciparum* detected, while a minority contained other species, mostly as mixed infections together with *P. falciparum* (Supplementary Table 1). In particular, 4.2 % (96) of positive slides contained mixed infections of *P. falciparum* and *P. malariae*, and 1.6 % (37) had mixed infection with *P. falciparum* and *P. ovale*. No mixed infections were detected with *P. malariae* and *P. ovale*, but a minority contained single species infections of either *P. malariae*, 1.7 % (38) and *P. ovale*, 1.5 % (33). As expected, no *P. vivax* or any zoonotic species was detected.

For all the positive slides except for eleven that had only *P. falciparum* gametocytes detected, the density of asexual malaria parasites was measured (Supplementary Table 1). Overall, 21.3% (2264) slides were positive for asexual malaria parasites, for which densities in blood ranged from 15 to 248185 μL^−1^, with a geometric mean of 2610 μL^−1^. Single-species infections with *P. falciparum* had a geometric mean density of 2671 μL^−1^, while the minority of single-species infections with *P. malariae* or *P. ovale* had respectively much lower geometric means of 933 μL^−1^ and 557 μL^−1^ (Supplementary Table 1). Mixed infections of *P. falciparum* with either of the other species had higher densities than for *P. falciparum* alone, a geometric mean of 4288 μL^−1^ for mixed *P. falciparum* and *P. ovale* infections, and 3355 μL^−1^ for mixed *P. falciparum* and *P. malariae* infections (Supplementary Table 1).

As *P. falciparum* comprised most of the malaria parasite infections, and these had much higher densities than the rarer species, the following analyses of parasite densities were performed on all infections together, and secondary analyses checked if any trends differed for *P. falciparum* alone.

### Malaria infection prevalence and asexual parasite density in different geopolitical zones of Nigeria

There was significant variation in prevalence of malaria parasites in children under five across the six major geopolitical zones in the country (P < 10^-15^) (Figure 1, Supplementary Table 2). This was attributable to the North-West zone having a much higher prevalence (31.4 %) than the other five zones, while these had similar prevalence to each other (between 16.5 % and 18.8 %) (Figure 1B, Supplementary Table 2). Moreover, asexual parasite density in positive slides showed further differences among the geopolitical zones (P = 0.0026) (Figure 1C, Supplementary Table 2). The North-West zone had the highest geometric mean density (3291 μL^−1^), but very high densities were also seen in the North-East (geometric mean of 3059 μL^−1^) and the South-West (geometric mean of 3262 μL^−1^).

There was no significant difference between male and female children for either the malaria parasite prevalence or the density of asexual parasites in those who were positive (Supplementary Table 2). Slide positivity varied with age in years, being lowest in those under one year old and highest in those four years old (P < 10^-15^) (Supplementary Table 2). The parasite densities in positive individuals did not differ significantly according to age in years (P = 0.080), although those under one year old had slightly lower density infections than other children up to five years of age (Supplementary Table 2).

### Malaria infection prevalence and asexual parasite density in all individual states of Nigeria

The variation in prevalence and parasite density was examined in all 37 of the individual states across the country including the Federal Capital Territory (Figure 2A). Asexual parasite prevalence varied widely among states, from 2.2% in Lagos State (South-West zone) to 45.3% in Zamfara State (North-West zone) (P < 10^-15^), reflecting substantial variation within each geopolitical zone (Figure A, Supplementary Table 2). Geometric mean parasite densities in positive slides also varied significantly among states (P = 0.0046), from 716 μL^−1^ in Akwa Ibom state (South-South zone) to 4877 μL^−1^ in Yobe State (North-East zone), again reflecting variation within each geopolitical zone (Figure 2B, Supplementary Table 2).

**Figure 2.**
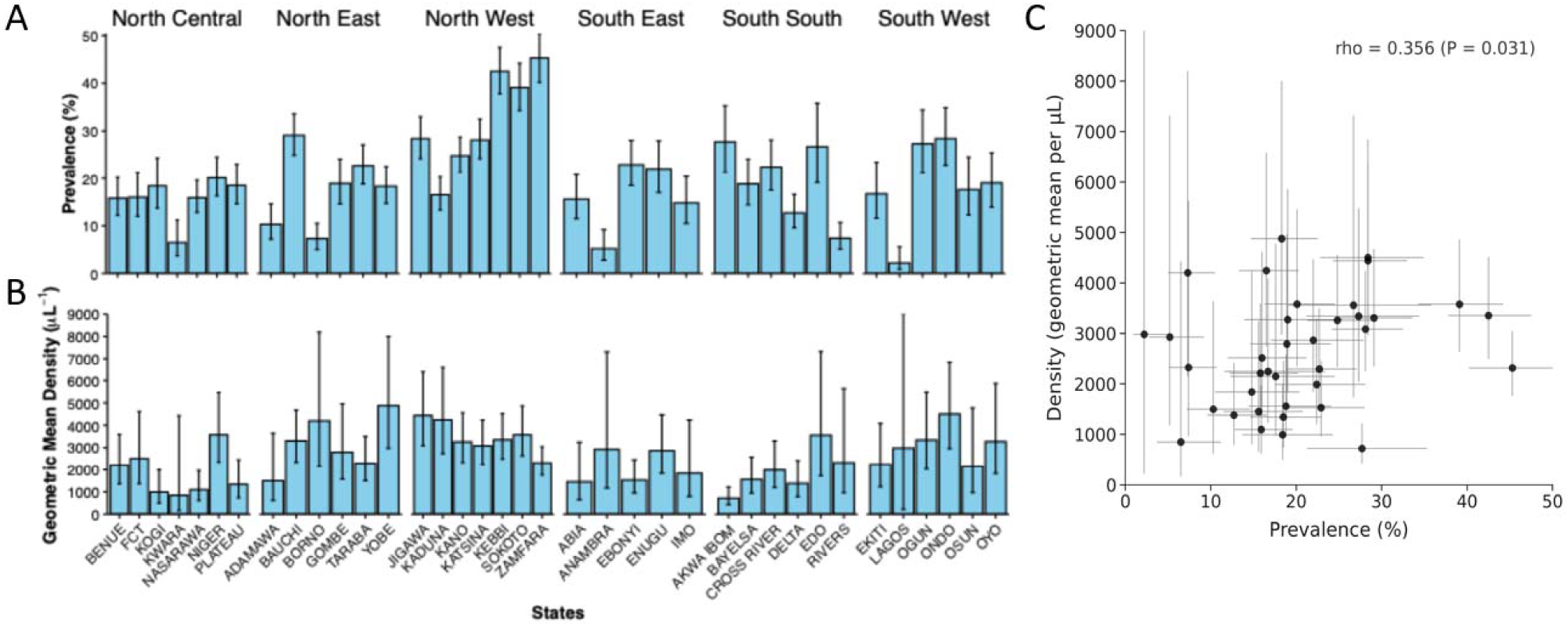
Asexual malaria parasite prevalence and density in children under five years of age in Nigeria, 2021. Individual states are grouped by geopolitical zone. **A.** Prevalence of asexual malaria parasite infection in each state, with 95% confidence intervals. Sample sizes of numbers of slides examined for each state (mean of 288) are shown in Supplementary Table 3. **B.** Geometric mean asexual parasite density in positive slides in each state, with 95% confidence intervals. Numbers of positive slides from which the densities were determined for each state are shown in Supplementary Table 2. The overall geometric mean density was 2610 µL^-1^ and values for each state are presented in Supplementary Table 3. **C.** Correlation between slide positivity prevalence and geometric mean parasite density in positive slides across all individual states (Spearman’s rho = 0.356, P = 0.031). Each point represents a state with horizontal and vertical bars indicating 95% confidence intervals.

Across all states, there was a significant correlation between the prevalence of parasite slide positivity and the geometric mean density of parasites in positive slides (Spearman’s rho *=* 0.356, P *=* 0.031) (Figure 2C). Most states in the North-West had high prevalence of infection as well as high parasite densities in those infected, emphasising that this zone clearly has the highest malaria infection burden within Nigeria.

### Sexual stage parasite prevalence in geopolitical zones and individual states of Nigeria

A total of 604 (5.7 %) of the 10651 children were positive for sexual stage malaria parasites, the gametocytes required for transmission. All except nine of these also had asexual parasites detected in the blood at the same time. Across all states, the prevalence of detectable sexual stage parasites correlated strongly with that of asexual parasites (Spearman’s rho = 0.869, P < 0.001) while being approximately four-fold lower on average (Figure 3A, Supplementary Table 3). The prevalence of sexual stage parasites was significantly higher in the North-West zone than in the rest of the country (Figure 3B, Supplementary Table 4), as was the case for asexual stage parasites described above.

**Figure 3.**
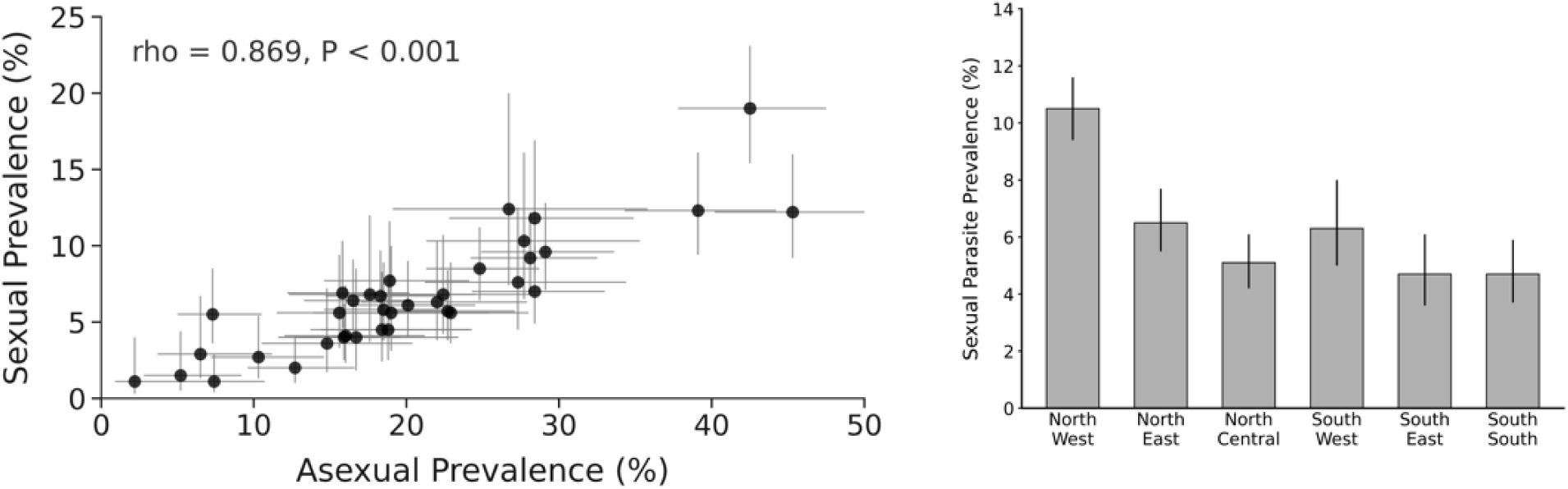
Sexual stage malaria parasite prevalence in Nigeria shows significant geographical variation. **A.** Correlation between prevalence of sexual and asexual malaria parasites in children under five years of age across all states in Nigeria (Spearman’s ρ = 0.869, *p* < 0.001). Each state is plotted as a point with bars indicating 95% confidence intervals for each prevalence estimate. Numbers for each state are shown in Supplementary Table 3. **B.** The sexual-stage parasite prevalence (shown with 95% confidence intervals) is much higher in the North-West than in each of the other geopolitical zones in Nigeria. The sexual-stage parasite prevalence is slightly higher in the North-East and South-West than in the remaining zones.

### Changes in malaria parasite indices in Nigeria between surveys in 2018 and 2021

Previous analysis of asexual parasite densities, presence of gametocytes and parasite species in slides collected from the 2018 national NDHS survey used similar sampling and laboratory methods ^18^, which enables direct comparisons with results obtained here from the 2021 survey slides. The 2021 MIS survey report had shown no significant difference in the overall slide positivity in 2021 compared to 2018, which initially suggested that there were no notable changes. However, more detailed analysis enabled by the data here indicates several significant changes.

Although the overall proportion of malaria parasite prevalence was similar in slides examined here from both surveys (21.5% in 2018 and 21.3% in 2021), the overall geometric mean asexual parasite density in positive slides was significantly higher in 2021 (2610 µL^-1^) than in 2018 (2241 µL^-1^). This varied across the country. The North-Central zone had a decrease in prevalence from 20.7% to 16.5% (P = 0.002) (Figure 4A, Supplementary Table 5) as well as a decrease in geometric mean asexual parasite density from 2535 to 1748 µL^-1^ in the positive slides (P = 0.053) (Figure 4B, Supplementary Table 5). No other zone showed a change in prevalence, but there were significant increases in parasite densities in the North-West (P = 0.022), North-East (P = 0.006), and South-West (P = 0.0001) in 2021 compared to 2018 (Figure 4B, Supplementary Table 5).

**Figure 4.**
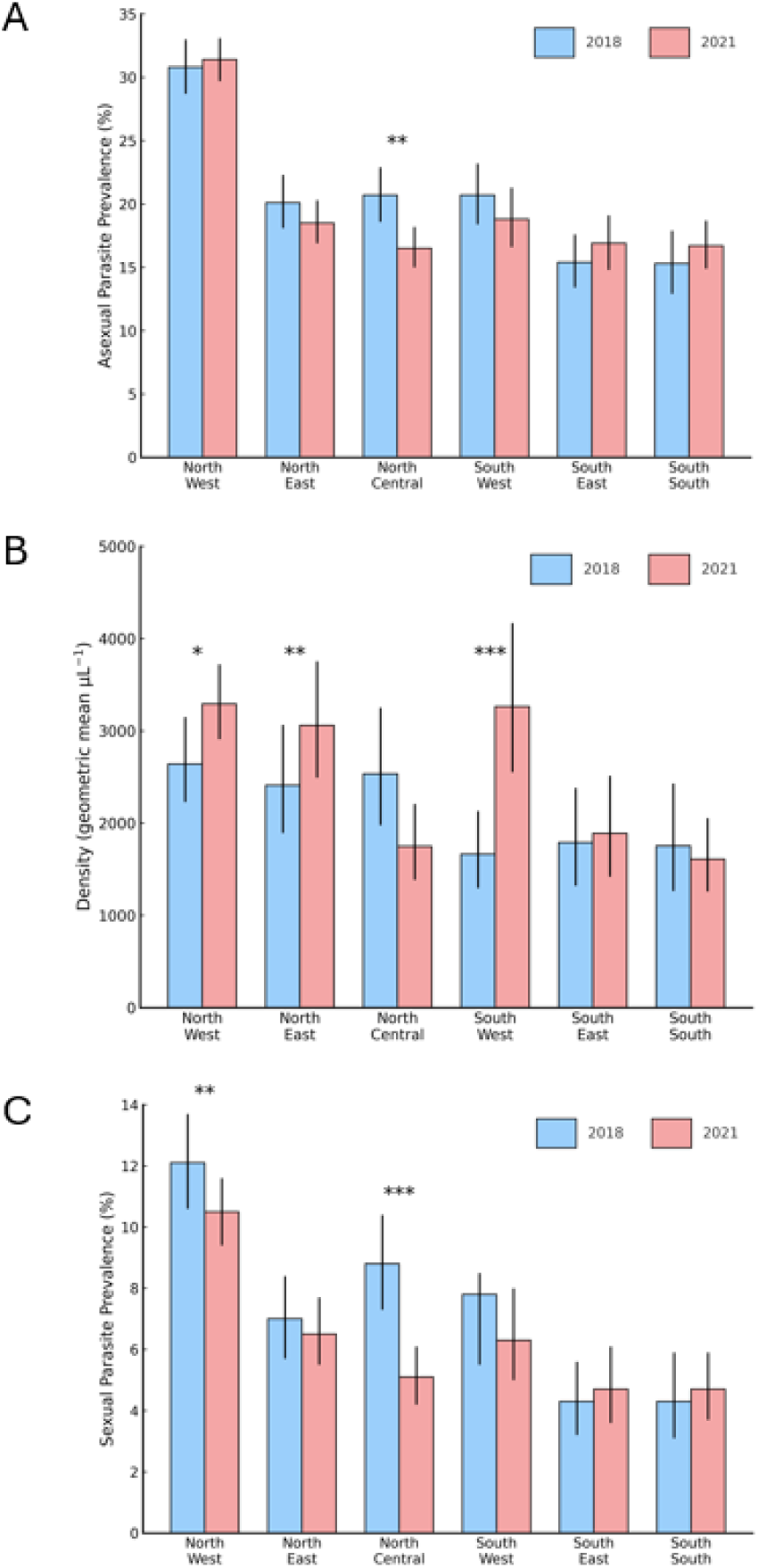
Comparisons between surveys in 2018 and 2021 of malaria parasite prevalence and density within infections in each of the geopolitical zones of Nigeria. **A.** Significant change in asexual parasite prevalence between 2018 and 2021 was seen in one zone (North-Central, P = 0.002). **B.** Significant increases in asexual parasite density within infections was seen in three zones (North-West, P = 0.022; North-East, P = 0.006; South-West, P = 0.0001). **C.** Significant reduction in sexual stage parasite prevalence was seen in two zones (from 12.1% to 10.5% in the North-West, P = 0.0012; from 8.8% to 5.1% in the North-Central zone, P < 10^-6^). All prevalence numbers and P values of tests are shown in Supplementary Tables 4 and 5.

Significant changes in the prevalence of sexual stage parasites between 2018 and 2021 were seen in two geopolitical zones, but not in the other four. In the North-Central zone there was a reduction in sexual stage parasite prevalence from 8.8% to 5.1% (P < 10^-6^), while in the North-West zone there was a reduction from 12.1% to 10.5% (P = 0.0012) (Figure 4C, Supplementary Table 4).

Comparing between 2018 and 2021 for each of the 37 individual states within the country including the Federal Capital Territory, there were significant differences in asexual parasite prevalence (at the P < 0.01 level for each comparison) in six states, lower for 2021 in five and higher for 2021 in one (Supplementary Dataset 1). There were significant differences in asexual parasite densities within infections in three other states, lower for 2021 in one and higher for 2021 in two (Supplementary Dataset 1). There were significant differences in sexual parasite prevalence in three states, lower for 2021 in two and higher for 2021 in one (Supplementary Dataset 1).

As in the case of the 2021 survey, most of the positive slides in 2018 contained *P. falciparum* alone (Figure 5, Supplementary Table 6) ^18^. Analysis of parasite densities in all positive slides showed that only *P. falciparum* infections had a higher geometric mean density in 2021 (2671 µL^-1^) compared with 2018 (2304 µL^-1^)(P = 0.0076), and there were no differences between the surveys in the densities of the other infections (Figure 5, Supplementary Table 6). Although a minority, it is notable that the prevalence of slides positive with *P. malariae* alone decreased significantly from 0.9 % in 2018 to 0.4 % in 2021 (P < 10^-6^). In parallel to this, the prevalence of mixed infections containing both *P. falciparum* and *P. malariae* decreased from 1.4 % to 0.9 % (P = 0.002).

**Figure 5.**
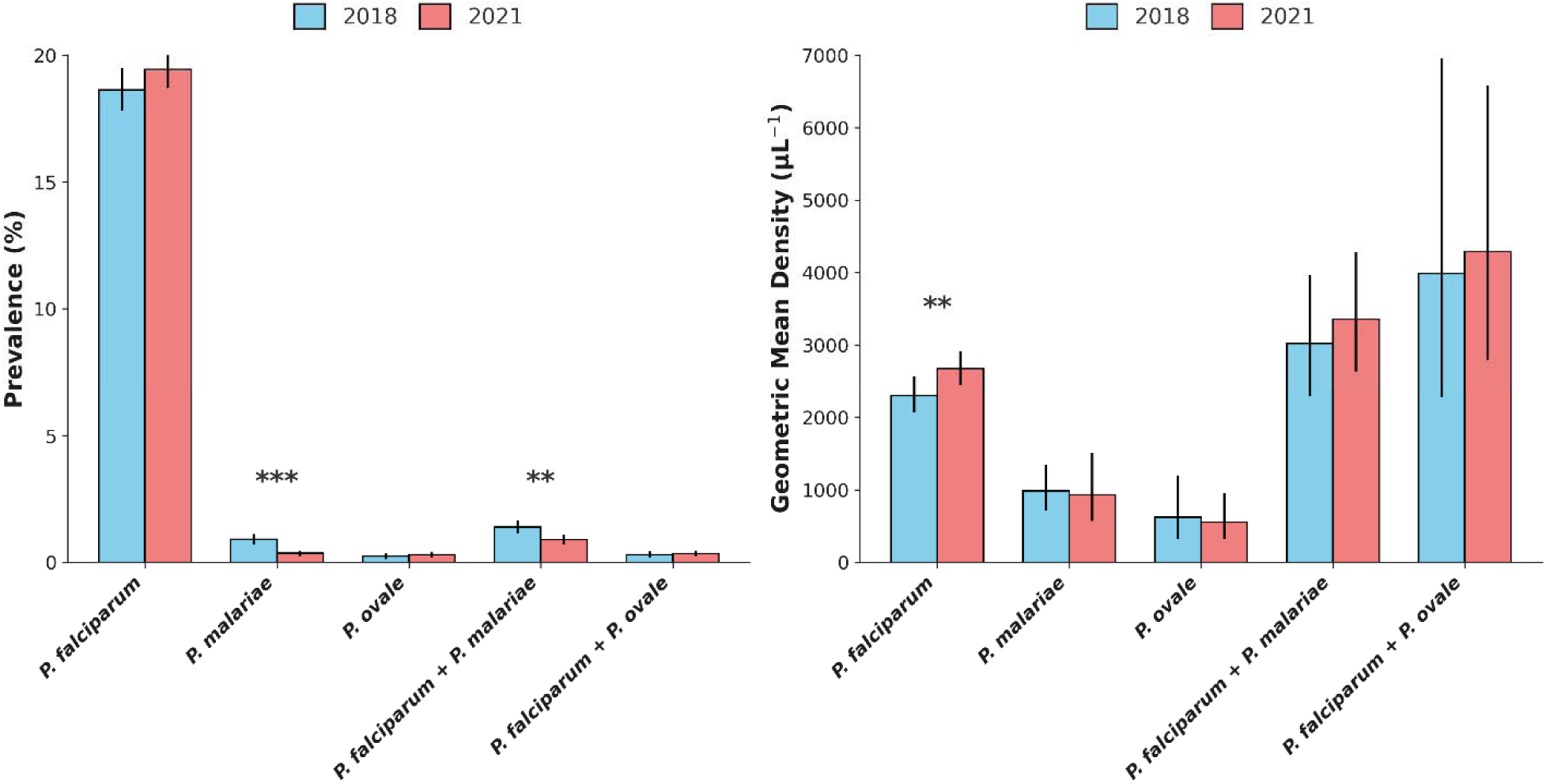
Comparison of asexual parasite prevalence and density for different *Plasmodium* species in Nigeria in the 2018 and 2021 surveys. **A.** Most positive slides in 2021 contained *P. falciparum* alone, and this was similar in the survey in 2018. Although a minority, the prevalence of slides positive with *P. malariae* alone decreased significantly from 0.9 % in 2018 to 0.4 % in 2021 (P < 10^-6^), and the prevalence of mixed infections containing both *P. falciparum* and *P. malariae* also decreased from 1.4 % to 0.9 % (P = 0.002). **B.** Analysis of parasite densities in all positive slides showed that only *P. falciparum* infections had a higher geometric mean density in 2021 (2671 µL^-1^) compared with 2018 (2304 µL^-1^)(P = 0.0076). There were no differences between the surveys in the densities of the other types of infections. Numerical values are presented in Supplementary Table 6.

## Discussion

This study on the largest nationwide survey of malaria shows that geographical and temporal trends are enhanced by asexual parasite density measurements and detection of sexual parasite transmission stages. Importantly, parasite prevalence and parasite density within infections are separate indices with different denominators, enabling independent analyses. The parasite density distributions show more geographical and temporal variation than revealed by comparisons of prevalence alone. The sexual parasite prevalence correlates with the asexual parasite prevalence, since it is biologically causally linked, but its measurement has potential to increase resolution of geographical and temporal trends affecting transmission ^22^.

These findings are relevant for understanding the heterogeneous and changing epidemiology of malaria within Nigeria. Previously, the three northern geopolitical zones of the country had higher prevalence of infection than southern zones ^17^ ^18^. In this 2021 survey, only the North-West had higher asexual parasite prevalence than other zones, but three zones (North-West, North-East, and South-West) had higher geometric mean parasite densities, and these densities were also higher than seen in the 2018 survey. Only one zone (North-Central) zone had a lower asexual parasite prevalence in 2021 than in 2018, but two zones (North-Central and North-West) had lower sexual parasite prevalence. Finer understanding of trends across the country was extended by examination of the changes within individual states, which showed clear variation within as well as between the major geopolitical zones.

More than 90% of all malaria infections in children under five in Nigeria contained *P. falciparum* alone. These had much higher parasite densities than single-species infections with the relatively uncommon *P. malariae* or *P. ovale* alone, while mixed-species infections of *P. falciparum* with either of the other parasites had higher densities than those with *P. falciparum* alone. The higher mean parasite density in 2021 compared with 2018 is specifically due to *P. falciparum*, and there were no changes in the density distributions of other malaria infections. A decrease in prevalence of *P. malariae* between 2018 and 2021 is notable, although it does not contribute to the major trends associated with *P. falciparum* here. *P. malariae* and *P. ovale* used to be more commonly detected by microscopy several decades ago than they have been in recent years ^23^. More sensitive detection of infections by species-specific PCR methods indicate that low-density infections with these species can persist for many years so that their prevalence may be more resilient to short-term control interventions compared with *P. falciparum* ^24^ ^25^. However, wherever *P. falciparum* incidence has declined over a long period these other species are now rarely detected, and they have not impeded the prospect of malaria elimination anywhere.

When expertise and facilities exist, there are strong reasons to use additional microscopy measurements in national surveys, as illustrated here. The methods are standardised and operate under WHO guidelines on accreditation and quality control. Moreover, results are comparable between different surveys over time, which is not the case for lateral-flow RDT results which are subject to variation in availability and performance of commercially available kits ^26^. Notably, there are only very marginal resource costs to obtain the additional microscopy measurements, on top of the standard scoring of parasite positivity. Such costs are manageable within countries with relevant expertise and facilities, and there is a high within-country motivation as the results deliver extra malaria indicators. As the future of international and domestic funding of national surveys is considered, this provides an evidence-based case for their inclusion.

Future surveys may also consider options of further methods in addition to expert microscopy. For example, qPCR assays are more sensitive to detect and estimate numbers of parasites in low density infections, which would yield higher prevalence estimates and extend the analysis of parasite density distributions ^9^ ^27^. Other detection methods such as loop-mediated isothermal amplification may also be more sensitive than microscopy ^28^, but do not yield estimates of parasite densities. An issue with these tests is that the protocols are not yet subject to WHO-recommended standardisation and laboratory accreditation for operation. Beyond the potential for parasite detection, parasite DNA-based surveillance offers ability to compare the genetic complexity of infections, and the distribution of drug resistance variants in different populations, which in Nigeria has so far been mostly limited to looking within clinical samples ^29^. It is also important to emphasise that the representative sampling here underpins the value of these population-based measures, recognising that variation in parasite densities and detection of gametocytes or alternative species would be expected among blood samples taken at different times from individuals ^30^ ^31^.

It is now recognised that long-term sustainability of large scale surveys requires increased agency and funding from malaria-endemic countries. A UN task force has recommended that countries take greater ownership by designing and funding their own surveys tailored to the information they need for policymaking, which supports a broader agenda for national leadership ^32^. Indeed, it has been recognised that a global funding crisis should serve as a ‘tipping point’ for these vital surveys, so that national governments and partners develop sustainable initiatives, while preserving the principles and quality structures developed by the DHS Program ^6^. By focusing on an important survey indicator that is relevant to the country with the highest malaria burden, the present study offers a significant contribution to this process.

## Author Contributions

All authors contributed to the study conduct; W.O., A.M.B., and D.J.C. led the analytical design; W.O., O.Ol., N.O., R.O.U.; G.O., C.An., G.N., C.O., P.U., N.E., O.Or, A.M., O.Ok., B.A., S.I., F.O., I.K., L.O., I.M., K.M., S.E., E.N., C.Am., M.A., O.M., Z.B., T.A., N.O., J.I., and A.L. were responsible for sample collection and survey data; W.O. supervised the generation and validation of laboratory slide microscopy data; W.O., A.M.B., D.J.C. performed data analyses; W.O., A.M.B., and D.J.C. wrote the manuscript; all authors considered and commented on manuscript drafts and approved the final version of the manuscript.

## Data Availability

All data generated during this study are included in this published article and its supplementary information files. All other data information from the 2021 Nigeria Malaria Indicator Survey are publicly available from the DHS Program (www.dhsprogram.com).

## References

1. WHO. World Malaria Report 2025. Geneva: World Health Organization, 2025.

2. WHO. Countries and territories certified malaria-free by WHO Geneva: WHO; 2026 [Available from: https://www.who.int/teams/global-malaria-programme/elimination/countries-and-territories-certified-malaria-free-by-who accessed July 3 2026.

3. Abeles J, Conway DJ. The Gini coefficient as a useful measure of malaria inequality among populations. Malar J 2020;19(1):444. doi: 10.1186/s12936-020-03489-x

4. Corsi DJ, Neuman M, Finlay JE, et al. Demographic and health surveys: a profile. Int J Epidemiol 2012;41(6):1602–13. doi: 10.1093/ije/dys184

5. Wahl B, Menon GI, Nyhan K, et al. Global health data precarity: Safeguarding the Demographic and Health Survey program as a global public good. Sci Data 2025;12(1):1827. doi: 10.1038/s41597-025-06128-9

6. Khaki JJ, Molenaar J, Karki S, et al. When health data go dark: the importance of the DHS Program and imagining its future. BMC Med 2025;23(1):241. doi: 10.1186/s12916-025-04062-6

7. Weiss DJ, Lucas TCD, Nguyen M, et al. Mapping the global prevalence, incidence, and mortality of Plasmodium falciparum, 2000-17: a spatial and temporal modelling study. Lancet 2019;394(10195):322–31. doi: 10.1016/S0140-6736(19)31097-9

8. Imwong M, Stepniewska K, Tripura R, et al. Numerical distributions of parasite densities during asymptomatic malaria. J Infect Dis 2016;213(8):1322–9. doi: 10.1093/infdis/jiv596

9. Whittaker C, Slater H, Nash R, et al. Global patterns of submicroscopic *Plasmodium falciparum* malaria infection: insights from a systematic review and meta-analysis of population surveys. Lancet Microbe 2021;2(8):e366–e74. doi: 10.1016/S2666-5247(21)00055-0

10. Mappin B, Cameron E, Dalrymple U, et al. Standardizing *Plasmodium falciparum* infection prevalence measured via microscopy versus rapid diagnostic test. Malar J 2015;14:460. doi: 10.1186/s12936-015-0984-9

11. Dhorda M, Ba EH, Kevin Baird J, et al. Towards harmonization of microscopy methods for malaria clinical research studies. Malar J 2020;19(1):324. doi: 10.1186/s12936-020-03352-z

12. Onyango L, Ouedraogo-Ametchie G, Ozodiegwu I, et al. Subnational tailoring of malaria interventions for strategic planning and prioritization: Experience and perspectives of five malaria programs. PLOS Glob Public Health 2025;5(5):e0003811. doi: 10.1371/journal.pgph.0003811

13. National Malaria Elimination Programme (NMEP) National Population Commission (NPopC) National Bureau of Statistics (NBS) and ICF International. Nigeria Malaria Indicator Survey 2015. Abuja, Nigeria, and Rockville, Maryland, USA NMEP, NPopC, and ICF International, 2016.

14. National Population Commission (NPC) [Nigeria] National Malaria Control Programme (NMCP) [Nigeria] and ICF International. Nigeria Malaria Indicator Survey 2010. Abuja, Nigeria: NPC, NMCP, and ICF International, 2012.

15. National Population Commission (NPC) [Nigeria] and ICF International. Nigeria Demographic and Health Survey 2018. Abuja, Nigeria, and Rockville, Maryland, USA: NPC and ICF, 2019.

16. National Malaria Elimination Programme (NMEP) [Nigeria] NPCNN, and ICF,. Nigeria Malaria Indicator Survey (MIS) 2021. Abuja, Nigeria, and Rockville, Maryland, USA NMEP, NPC, and ICF, 2022.

17. Oyibo W, Ntadom G, Uhomoibhi P, et al. Geographical and temporal variation in reduction of malaria infection among children under 5 years of age throughout Nigeria. BMJ Glob Health 2021;6(2) doi: 10.1136/bmjgh-2020-004250

18. Oyibo W, Latham V, Oladipo O, et al. Malaria parasite density and detailed qualitative microscopy enhances large-scale profiling of infection endemicity in Nigeria. Sci Rep 2023;13(1):1599. doi: 10.1038/s41598-023-27535-1

19. WHO. Collection of finger-prick blood and preparation of thick and thin blood films (SOP 05a) 2016 [Available from: https://www.who.int/publications/i/item/HTM-GMP-MM-SOP-05a (last accessed 9th October 2025).

20. WHO. Malaria Microscopy Quality Assurance Manual: Version 2. Geneva, 2016.

21. WHO. Malaria Parasite Counting (SOP09): World Health Organization, 2016.

22. Barry A, Bradley J, Stone W, et al. Higher gametocyte production and mosquito infectivity in chronic compared to incident *Plasmodium falciparum* infections. Nat Commun 2021;12(1):2443. doi: 10.1038/s41467-021-22573-7

23. Molineaux L, Gramiccia G. The Garki project: research on the epidemiology and control of malaria in the Sudan Savanna of West Africa: World Health Organization 1980.

24. Yman V, Wandell G, Mutemi DD, et al. Persistent transmission of *Plasmodium malariae* and *Plasmodium ovale* species in an area of declining *Plasmodium falciparum* transmission in eastern Tanzania. PLoS Negl Trop Dis 2019;13(5):e0007414. doi: 10.1371/journal.pntd.0007414

25. Sutherland CJ. Persistent parasitism: The adaptive biology of malariae and ovale malaria. Trends Parasitol 2016;32(10):808–19. doi: 10.1016/j.pt.2016.07.001

26. Kavanaugh MJ, Azzam SE, Rockabrand DM. Malaria rapid diagnostic tests: Literary review and recommendation for a quality assurance, quality control algorithm. Diagnostics 2021;11(5) doi: 10.3390/diagnostics11050768

27. Slater HC, Ross A, Felger I, et al. The temporal dynamics and infectiousness of subpatent *Plasmodium falciparum* infections in relation to parasite density. Nat Commun 2019;10(1):1433. doi: 10.1038/s41467-019-09441-1

28. Rakotomalala Robinson D, Pennisi I, Cavuto ML, et al. Sensitive near point-of-care detection of asymptomatic and submicroscopic *Plasmodium falciparum* infections in African endemic countries. Nat Commun 2025;16(1):8925. doi: 10.1038/s41467-025-64027-4

29. Ajibaye O, Oboh-Imafidon M, Ajibola O, et al. Molecular surveillance of malaria in Nigeria reveals a low frequency of antimalarial resistance markers and clonal expansion of chloroquine-sensitive infections. Open Forum Infect Dis 2026;13(1):ofaf719. doi: 10.1093/ofid/ofaf719

30. Farnert A, Lebbad M, Faraja L, et al. Extensive dynamics of *Plasmodium falciparum* densities, stages and genotyping profiles. Malar J 2008;7:241. doi: 10.1186/1475-2875-7-241

31. Andolina C, Ramjith J, Rek J, et al. *Plasmodium falciparum* gametocyte carriage in longitudinally monitored incident infections is associated with duration of infection and human host factors. Sci Rep 2023;13(1):7072. doi: 10.1038/s41598-023-33657-3

32. Nott J, Kaunda-Khangamwa B, Mathanga DP, et al. Beyond the Demographic and Health Survey: on the past and future of population health surveillance. BMJ Glob Health 2026;11(2) doi: 10.1136/bmjgh-2025-022023

