## Supplementary Table for "Increased sensitivity of detecting variation in malaria endemicity by parasite density measurement in large surveys"

**Supplementary Table 1.** Prevalence and asexual parasite density of infections by different malaria parasite species in children under five years of age in Nigeria in the 2021 nationwide survey.

|  | **Prevalence** | | **Density within infections** | |
| --- | --- | --- | --- | --- |
|  | **% (N = 10651)** | **95% CI** | **Geometric mean μL^−1^** | **95% CI** |
| *P. falciparum* | 19.34 (2060) | 18.70 – 20.22 | 2671 | 2447 - 2916 |
| *P. malariae* | 0.36 (38) | 0.24 - 0.47 | 933 | 573 - 1519 |
| *P. ovale* | 0.31 (33) | 0.20 – 0.42 | 557 | 323 - 960 |
| *P. falciparum* and  *P. malariae* | 0.90 (96) | 0.72 – 1.08 | 3355 | 2630 - 4279 |
| *P. falciparum* and  *P. ovale* | 0.35 (37) | 0.24 – 0.46 | 4288 | 2792 - 6585 |

**Supplementary Table 2.** Asexual malaria parasite prevalence and density in infected children under five years of age in Nigeria surveyed in 2021, tabulated by sex, age, and geopolitical zone.

|  | **Prevalence** | | | **Density** | | |
| --- | --- | --- | --- | --- | --- | --- |
|  | **Asexual parasite prevalence (%)** | **95% CI (%)** | **P-value^a^** | **Asexual parasite density in infected children (geometric mean μL^−1^)** | **95% CI (%)** | **P-value^b^** |
| **Sex** | | | | | | |
| **Male** | 21.2 (1158/5468) | 20.1 – 22.3 | 0.86 | 2588 | 2306 - 2905 | 0.90 |
| **Female** | 21.3 (1106/5183) | 20.2 – 22.5 |  | 2634 | 2343 -2960 |  |
| **Age (years)** | | | | | | |
| **<1** | 13.1 (128/975) | 11.2 – 15.4 | < 10^-15^ | 1800 | 1250 -2591 | 0.080 |
| **1** | 15.6 (319/2051) | 14.0 – 17.2 |  | 2537 | 2026 -3178 |  |
| **2** | 18.8 (424/2259) | 17.2 – 20.4 |  | 2948 | 2440 -3562 |  |
| **3** | 23.2 (567/2444) | 21.6 – 24.9 |  | 3110 | 2634 -3672 |  |
| **4** | 28.3 (826/2922) | 26.7 – 29.9 |  | 2329 | 2042 -2656 |  |
| **Geopolitical zone** | | | | | | |
| **North-Central** | 16.5 (342/2071) | 15.0 – 18.2 | < 10^-15^ | 1748 | 1384 -2208 | 0.0026 |
| **North-East** | 18.5 (378/2039) | 16.9 – 20.3 |  | 3059 | 2493 - 3754 |  |
| **North-West** | 31.4 (915/2918) | 29.7 – 33.1 |  | 3291 | 2910 - 3722 |  |
| **South-East** | 16.9 (193/1145) | 14.8 – 19.1 |  | 1889 | 1419 - 2514 |  |
| **South-South** | 16.7 (240/1438) | 14.9 – 18.7 |  | 1609 | 1259 - 2055 |  |
| **South-West** | 18.8 (196/1040) | 16.6 – 21.3 |  | 3262 | 2553 - 4169 |  |

*^a^Pearson’s chi-squared test. ^b^Mann-Whitney or Kruskall-Wallis tests.*

**Supplementary Table 3.** Prevalence of asexual malaria parasites and geometric mean asexual parasite densities within infections as well as prevalence of sexual stage malaria parasites in each state of Nigeria. The dataset comprises 10651 children up to five years of age among those in the 2021 MIS survey with complete microscopy measurements for relevant variables.

| **State** | **N** | **Asexual Parasite Prevalence** | | | **Asexual Parasite Density in positive slides** | | **Sexual Parasite Prevalence** | | |
| --- | --- | --- | --- | --- | --- | --- | --- | --- | --- |
|  |  | **Positive slides** | **Prevalence (%)** | **95% CI** | **Geometric Mean Density** | **95% CI** | **Positive slides** | **Prevalence (%)** | **95% CI** |
| ABIA | 231 | 36 | 15.6 | 11.5 - 20.8 | 1452 | 650 - 3246 | 13 | 5.6 | 3.3 - 9.4 |
| ADAMAWA | 261 | 27 | 10.3 | 7.2 - 14.6 | 1498 | 616 - 3643 | 7 | 2.7 | 1.3 - 5.4 |
| AKWA IBOM | 155 | 43 | 27.7 | 21.3 - 35.3 | 717 | 421 - 1220 | 16 | 10.3 | 6.5 - 16.1 |
| ANAMBRA | 194 | 10 | 5.2 | 2.8 - 9.2 | 2926 | 1171 - 7312 | 3 | 1.5 | 0.5 - 4.4 |
| BAUCHI | 416 | 121 | 29.1 | 24.9 - 33.6 | 3306 | 2339 - 4674 | 40 | 9.6 | 7.1 - 12.8 |
| BAYELSA | 245 | 46 | 18.8 | 14.4 - 24.1 | 1558 | 943 - 2576 | 11 | 4.5 | 2.5 - 7.9 |
| BENUE | 317 | 50 | 15.8 | 12.2 - 20.2 | 2212 | 1362 - 3594 | 22 | 6.9 | 4.6 - 10.3 |
| BORNO | 343 | 25 | 7.3 | 5 - 10.5 | 4200 | 2153 - 8194 | 19 | 5.5 | 3.6 - 8.5 |
| CROSS RIVER | 237 | 53 | 22.4 | 17.5 - 28.1 | 1987 | 1197 - 3300 | 16 | 6.8 | 4.2 - 10.7 |
| DELTA | 347 | 44 | 12.7 | 9.6 - 16.6 | 1380 | 789 - 2414 | 7 | 2 | 1 - 4.1 |
| EBONYI | 301 | 69 | 22.9 | 18.5 - 28 | 1528 | 951 - 2455 | 17 | 5.6 | 3.6 - 8.9 |
| EDO | 105 | 28 | 26.7 | 19.1 - 35.8 | 3559 | 1729 - 7325 | 13 | 12.4 | 7.4 - 20 |
| EKITI | 150 | 25 | 16.7 | 11.6 - 23.4 | 2245 | 1234 - 4085 | 6 | 4 | 1.8 - 8.5 |
| ENUGU | 223 | 49 | 22 | 17 - 27.9 | 2866 | 1839 - 4467 | 14 | 6.3 | 3.8 - 10.3 |
| FCT | 243 | 39 | 16 | 12 - 21.2 | 2515 | 1371 - 4616 | 10 | 4.1 | 2.3 - 7.4 |
| GOMBE | 259 | 49 | 18.9 | 14.6 - 24.1 | 2791 | 1571 - 4960 | 20 | 7.7 | 5.1 - 11.6 |
| IMO | 196 | 29 | 14.8 | 10.5 - 20.4 | 1840 | 800 - 4234 | 7 | 3.6 | 1.7 - 7.2 |
| JIGAWA | 398 | 113 | 28.4 | 24.3 - 33 | 4440 | 3089 - 6383 | 28 | 7 | 4.9 - 10 |
| KADUNA | 424 | 70 | 16.5 | 13.3 - 20.3 | 4244 | 2736 - 6583 | 27 | 6.4 | 4.4 - 9.1 |
| KANO | 520 | 129 | 24.8 | 21.3 - 28.7 | 3259 | 2331 - 4558 | 44 | 8.5 | 6.4 - 11.2 |
| KATSINA | 448 | 126 | 28.1 | 24.2 - 32.5 | 3083 | 2244 - 4236 | 41 | 9.2 | 6.8 - 12.2 |
| KEBBI | 395 | 168 | 42.5 | 37.8 - 47.5 | 3353 | 2490 - 4517 | 75 | 19 | 15.4 - 23.1 |
| KOGI | 201 | 37 | 18.4 | 13.7 - 24.3 | 992 | 493 - 1998 | 9 | 4.5 | 2.4 - 8.3 |
| KWARA | 170 | 11 | 6.5 | 3.7 - 11.2 | 846 | 162 - 4427 | 5 | 2.9 | 1.3 - 6.7 |
| LAGOS | 180 | 4 | 2.2 | 0.9 - 5.6 | 2980 | 219 - 40566 | 2 | 1.1 | 0.3 - 4 |
| NASARAWA | 452 | 72 | 15.9 | 12.8 - 19.6 | 1096 | 611 - 1965 | 18 | 4 | 2.5 - 6.2 |
| NIGER | 363 | 73 | 20.1 | 16.3 - 24.5 | 3580 | 2347 - 5462 | 22 | 6.1 | 4.0 - 9.0 |
| OGUN | 172 | 47 | 27.3 | 21.2 - 34.4 | 3342 | 2039 - 5478 | 13 | 7.6 | 4.5 - 12.5 |
| ONDO | 211 | 60 | 28.4 | 22.8 - 34.9 | 4502 | 2961 - 6844 | 25 | 11.8 | 8.2 - 16.9 |
| OSUN | 148 | 26 | 17.6 | 12.3 - 24.5 | 2148 | 966 - 4777 | 10 | 6.8 | 3.7 - 12 |
| OYO | 179 | 34 | 19 | 13.9 - 25.4 | 3273 | 1827 - 5863 | 10 | 5.6 | 3.1 - 10 |
| PLATEAU | 325 | 60 | 18.5 | 14.6 - 23 | 1343 | 734 - 2459 | 19 | 5.8 | 3.8 - 8.9 |
| RIVERS | 349 | 26 | 7.4 | 5.1 - 10.7 | 2325 | 959 - 5634 | 4 | 1.1 | 0.4 - 2.9 |
| SOKOTO | 373 | 146 | 39.1 | 34.3 - 44.2 | 3578 | 2633 - 4862 | 46 | 12.3 | 9.4 - 16.1 |
| TARABA | 388 | 88 | 22.7 | 18.8 - 27.1 | 2295 | 1506 - 3498 | 22 | 5.7 | 3.8 - 8.4 |
| YOBE | 372 | 68 | 18.3 | 14.7 - 22.5 | 4878 | 2973 - 8002 | 25 | 6.7 | 4.6 - 9.7 |
| ZAMFARA | 360 | 163 | 45.3 | 40.2 - 50.4 | 2314 | 1757 - 3046 | 44 | 12.2 | 9.2 - 16 |

**Supplementary Table 4.** Comparison of sexual-stage malaria parasite prevalence among children under five years of age in Nigeria in 2018 (previously reported <https://doi.org/10.1038/s41598-023-27535-1>) and 2021 (this study), by sex, age group, and geopolitical zone.

|  | **2018** | | | **2021** | | | **2018 vs 2021** |
| --- | --- | --- | --- | --- | --- | --- | --- |
|  | **Sexual parasite prevalence (%)** | **95% CI (%)** | **P-value**^a^ | **Sexual parasite prevalence** | **95% CI (%)** | **P-value**^a^ | **P-value**^a^ |
| **Sex** | | | | | | |  |
| **Male** | 8.2 (324/3971) | 7.3-9.1 | 0.20 | 6.9 (379/5468) | 6.3-7.6 | 0.77 | 0.0276 |
| **Female** | 7.3 (280/3812) | 6.5-8.2 |  | 6.8 (351/5183) | 6.1-7.5 |  | 0.313 |
| **Age (years)** | | | | | | |  |
| **<1** | 4.9 (37/717) | 3.5-6.7 | < 0.001 | 5.9 (58/975) | 4.6-7.6 | 0.0047 | 0.558 |
| **1** | 6.3 (103/1534) | 5.2-7.6 |  | 5.4 (110/2051) | 4.5-6.4 |  | 0.105 |
| **2** | 7.6 (124/1514) | 6.3-9.0 |  | 6.9 (155/2259) | 5.9-8.0 |  | 0.143 |
| **3** | 8.8 (154/1598) | 7.5-10.2 |  | 7.0 (172/2444) | 6.1-8.1 |  | 0.00363 |
| **4** | 9.7 (160/1493) | 8.3-11.2 |  | 8.0 (235/2922) | 7.1-9.1 |  | 0.00386 |
| **Geopolitical zone** | | | | | | |  |
| **North-Central** | 8.8 (120/1248) | 7.3-10.4 | < 0.001 | 5.1 (105/2071) | 4.2-6.1 | <10^-15^ | 0.00000066 |
| **North-East** | 7.0 (97/1294) | 5.7-8.4 |  | 6.5 (133/2039) | 5.5-7.7 |  | 0.312 |
| **North-West** | 12.1 (217/1579) | 10.6-13.7 |  | 10.5 (305/2918) | 9.4-11.6 |  | 0.0012 |
| **South-East** | 4.3 (52/1155) | 3.2-5.6 |  | 4.7 (54/1145) | 3.6-6.1 |  | 0.884 |
| **South-South** | 4.3 (37/815) | 3.1-5.9 |  | 4.7 (67/1438) | 3.7-5.9 |  | 0.980 |
| **South-West** | 7.8 (81/1087) | 5.5-8.5 |  | 6.3 (66/1040) | 5.0-8.0 |  | 0.358 |

^a^Chi-squared test

**Supplementary Table 5.** Comparison of asexual malaria parasite prevalence and geometric mean parasite density by geopolitical zone in Nigeria, between 2018 (previously reported <https://doi.org/10.1038/s41598-023-27535-1>) and 2021 (this study).

**A. Parasite Prevalence**

| **Zone** | **2018** | | **2021** | | **P value ^a^** |
| --- | --- | --- | --- | --- | --- |
|  | **Prevalence (%)** | **95% CI (%)** | **Prevalence (%)** | **95% CI (%)** |  |
| **North-Central** | 20.7 | 18.6 – 22.9 | 16.5 | 15.0 – 18.2 | 0.00221 |
| **North-East** | 20.1 | 18.1 – 22.3 | 18.5 | 16.9 – 20.3 | 0.264 |
| **North-West** | 30.8 | 28.7 – 33.0 | 31.4 | 29.7 – 33.1 | 0.728 |
| **South-East** | 15.4 | 13.4 – 17.6 | 16.9 | 14.8 – 19.1 | 0.370 |
| **South-South** | 15.3 | 12.9 – 17.9 | 16.7 | 14.9 – 18.7 | 0.400 |
| **South-West** | 20.7 | 18.4 – 23.2 | 18.8 | 16.6 – 21.3 | 0.295 |

**B. Parasite Density**

| **Zone** | **2018** | | **2021** | | **P value ^b^** |
| --- | --- | --- | --- | --- | --- |
|  | **Density (geometric mean μL^−1^)** | **95% CI (%)** | **Density (geometric mean μL^−1^)** | **95% CI (%)** |  |
| **North-Central** | 2535 | 1977 – 3251 | 1748 | 1384 -2208 | 0.0527 |
| **North-East** | 2409 | 1893 – 3065 | 3059 | 2493 - 3754 | 0.0056 |
| **North-West** | 2640 | 2229 –3151 | 3291 | 2910 - 3722 | 0.0218 |
| **South-East** | 1789 | 1322 – 2382 | 1889 | 1419 - 2514 | 0.586 |
| **South-South** | 1752 | 1264 – 2429 | 1609 | 1259 - 2055 | 0.853 |
| **South-West** | 1663 | 1296 – 2133 | 3262 | 2553 - 4169 | 0.00013 |

^a^Pearson’s chi-squared test. ^b^Mann-Whitney test.

**Supplementary Table 6.** Comparison of prevalence and geometric mean density of different malaria parasite species in Nigeria in 2018 (previously reported <https://doi.org/10.1038/s41598-023-27535-1>) and 2021 (this study), showing single and mixed-species infections separately.

| **Parasite Species** | | **Prevalence (%, with 95% confidence intervals and numerators)** | | | **Parasite density (μL^−1^,** g**eometric mean with 95% CI )** | | |
| --- | --- | --- | --- | --- | --- | --- | --- |
|  |  | **2018**  **(N = 7783)** | **2021**  **(N = 10651)** | **P value^a^** | **2018** | **2021** | **P value^b^** |
| **Single** | *P. falciparum* | 18.63  (17.8-19.5)  (1450) | 19.44  (18.7-20.2)  (2071) | 0.17 | 2304  (2066-2570) | 2671  (2447-2916) | 0.0076 |
|  | *P. malariae* | 0.91  (0.70-1.12)  (71) | 0.36  0.24-0.47)  (38) | < 10^-5^ | 982  (718-1343) | 933  (573-1519) | 0.83 |
|  | *P. ovale* | 0.24  (0.13-0.35)  (19) | 0.31  (0.20-0.42) (33) | 0.49 | 622  (322-1202) | 557  (323-960) | 0.82 |
| **Mixed** | *P. falciparum & P. malariae* | 1.40  (1.14-1.66) (109) | 0.90  (0.72-1.08)  (96) | 0.0018 | 3021  (2298-3971) | 3355  (2630-4279) | 0.63 |
|  | *P. falciparum & P. ovale* | 0.31  (0.20-0.45)  (24) | 0.35  (0.24-0.46)  (37) | 0.75 | 3984  (2282-6955) | 4288  (2792-6585) | 0.84 |

Prevalence is shown as a percentage with absolute counts in parentheses. Geometric mean parasite density is calculated for positive individuals only. ^a^ Pearson’s chi-squared test. ^b^ Mann-Whitney test. Data from 2018 are previously published (Oyibo et al., Scientific Reports 2023) while data from 2021 are from the present study.
